# Economic cost of suicide of culturally and linguistically diverse populations in Australia

**DOI:** 10.1101/2024.07.18.24310624

**Authors:** Humaira Maheen, Chris Doran

## Abstract

**Background:** Suicide and self-harm pose significant global public health challenges and have substantial economic implications. Recent evidence from Australia has indicated significant variations in the prevalence of suicidal behaviours and mortality among diverse populations. Aim: This study aims to examine the economic cost of suicide among culturally and linguistically diverse (CALD) migrants in Australia.

**Methods:** We evaluated the economic impact of suicide by considering years of life lost, years of productive life lost, and overall economic costs, including direct, indirect, and intangible costs. We used data on suicide deaths in 2020 from the National Coroners Information System.

**Results:** The estimated annual economic cost of these suicides is $2.9 billion (in 2023 dollars). The average cost per suicide is $8.04 million for males and $9.23 million for females. The value of a statistical life year is the most significant cost driver, representing 94% of the total economic burden.

**Limitation:** These estimates do not capture costs associated with suicidal ideation, thoughts or self-harm attempts, which may substantially increase the economic burden.

**Conclusion:** This study emphasises the significant economic impacts of CALD suicide in Australia and highlights the urgent need for a comprehensive national suicide prevention program tailored for diverse populations.

## Introduction

Suicide is a public health concern worldwide. In the Global North, home to the most industrialised countries, it remains a leading cause of death (World Population Review, 2024). Current evidence from these countries has identified modifiable risk factors associated with suicide and suicidal behaviours (Franklin *et al*., 2017) along with effective prevention (Zalsman *et al.,* 2016) and postvention strategies (Andriessen *et al*., 2019). Over the past two decades, some of the countries in this region have seen a significant influx of economic and refugee migrants due to geopolitical conflicts, climate change, and ongoing economic uncertainty (OECD, 2023). Yet, suicide prevention efforts there often overlook stressors specific to migrant populations, resulting in missed opportunities for targeted initiatives. This oversight is particularly concerning for migrants from diverse cultural or ethnic backgrounds who differ from the local population in terms of race, ethnicity, language, and cultural beliefs about mental health and help-seeking practices(Forte *et al*., 2018; Chu *et al*., 2011; Mihtsintu *et al*., 2023). As a consequence, local prevention initiatives may not be relatable or effective for them.

Calculating the economic cost of suicide may seem morbid and morally inappropriate, yet it is one of the most effective ways to draw policymakers’ attention to this critical issue (McDaid, 2009). The economic impact of suicide includes not only the direct costs associated with the individual’s death and the indirect cost of the productive years the individual could have contributed to society but also the significant intangible nature of the psychological distress experienced by their friends and family members. Evidence from various developed countries has noted the significant economic cost of suicide, including in the United States ($53.1 billion in 2015) (Shepard, 2016), France (€18.5 billion in 2019) (Segar *et al*., 2024), and Ireland (€835 million in 2002) (Kennelly, 2007). In Australia, the total economic cost of suicidal behaviour (including direct, indirect and intangible costs) was recently estimated to be in the order of AUD 30.5 billion in 2018 (Productivity Commission, 2020). The price is substantially high in some high-risk groups, e.g., young people (AUD 511 million in 2014) (Kinchin & Doran, 2017) or construction industry workers (AUD 3.97 billion in 2019) (Productivity Commission, 2020; Doran & Potts, 2024).

In Australia, nearly 3,000 people die by suicide annually, and around 65,000 individuals make a suicide attempt every year (Australian Institute of Health and Welfare, 2023). In addition to the profound personal loss, suicide deaths affect up to 135 people, including family members, work colleagues, friends, and first responders present at the time of death (Cerel *et al*., 2019). Australia’s multicultural landscape, where approximately 30% of the population comes from culturally and linguistically diverse (CALD) backgrounds, adds complexity to this challenge. Recent evidence on suicidal behaviours amongst diverse populations noted significant differences between diverse groups on the prevalence of suicidal thoughts, attempts, and mortality (Maheen *et al*., 2024; Maheen & King, 2023). For instance, Pasifika migrants (men and women) have higher suicide rates compared to other migrant groups-which remain consistent over time, or African female migrants, whose suicide rates have increased by 8% between 2006-2019 (Maheen & King, 2023). As the demographic composition of Australia continues to evolve, there is a pressing need for government investment in tailored suicide prevention programs for CALD migrants. Currently, there is a lack of evidence regarding the economic cost of suicide among CALD migrants. This paper aims to address this gap by providing a comprehensive evaluation of the economic cost of suicide among CALD migrants in Australia.

## Methods

The impact of suicide among CALD migrants was assessed using three measures: (1) years of life lost (YLL), (2) years of productive life lost (YPLL), and (3) the economic cost, which includes direct, indirect, and intangible costs.

### Suicide data

Data on suicide deaths was obtained from the National Coroners Information System (NCIS), a web-based repository of coronial data from Australia and New Zealand. The NCIS holds online records of coronial briefs created as part of investigations conducted by coroners into individual deaths. Restricted access is granted to academics and researchers for research purposes after ethics approval. For this study, we obtained data on age, sex, employment status, country of birth, and usual country of residence for individuals who died by suicide in 2020. This information was only extracted for ‘closed’ cases, with complete coronial records.

### Definition of CALD population

For the purpose of this study, we used country of birth and usual country of residence as proxy indicators of CALD migrants. The NCIS uses the Standard Australian Classification of Countries (SACC) to code countries (Australian Bureau of Statistics, 2016). To identify cases from CALD backgrounds, we included those who were born in non-English speaking countries as defined by the Australian Bureau of Statistics (2013) and where Australia was noted as the usual country of residence. This excludes individuals born in Australia, Canada, New Zealand, Ireland, the United States, South Africa, the UK, and Ireland. Using a country of birth or corresponding geographical region is considered an acceptable approach and is particularly useful for studying migrant populations (Khan *et al*., 2022) and has also been used in the peer-reviewed literature (Maheen & King, 2023; Ide *et al*., 2012). Table 1 outlines the inclusion and exclusion criteria used to identify individuals from CALD populations for the study.

**Table 1:**
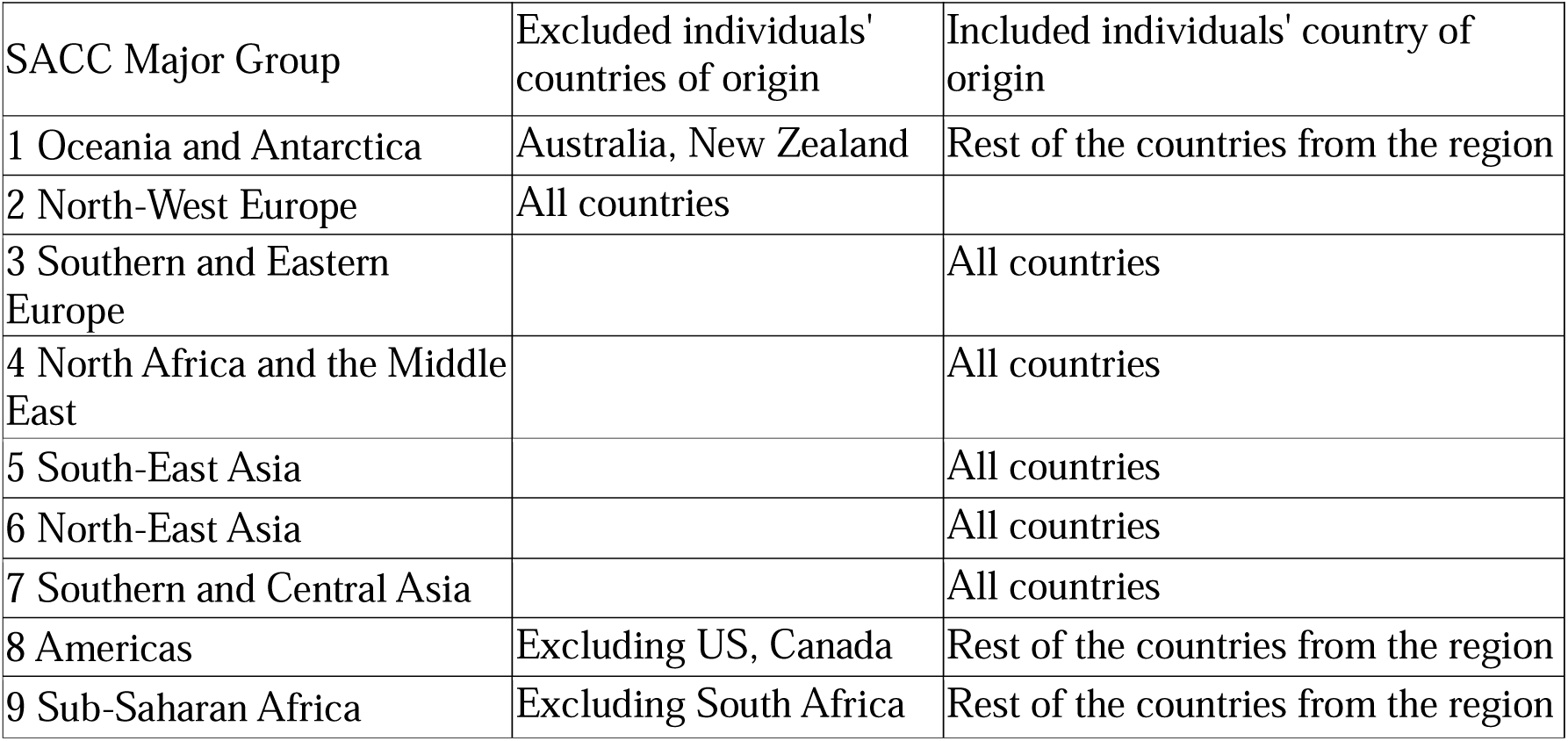
Inclusion and exclusion criterion.

| SACC Major Group | Excluded individuals' countries of origin | Included individuals' country of origin |
| --- | --- | --- |
| 1 Oceania and Antarctica | Australia, New Zealand | Rest of the countries from the region |
| 2 North-West Europe | All countries |  |
| 3 Southern and Eastern Europe |  | All countries |
| 4 North Africa and the Middle East |  | All countries |
| 5 South-East Asia |  | All countries |
| 6 North-East Asia |  | All countries |
| 7 Southern and Central Asia |  | All countries |
| 8 Americas | Excluding US, Canada | Rest of the countries from the region |
| 9 Sub-Saharan Africa | Excluding South Africa | Rest of the countries from the region |

#### 1. Average years of life lost (YLL)

The average age of a death by suicide (from NCIS) was subtracted from the average life expectancy at birth, 85.8 years for females and 83.2 years for males, to obtain an estimate of average YLL per fatality (United Nations Department of Economic and Social Affairs Population Division, 2022).

#### 2. Years of productive life lost (YPLL)

The average YPLL was derived by subtracting the average age of a death by suicide (from NCIS) from the retirement age in Australia (66.5 years) (Department of Social Services, 2016).

#### 3. Economic cost

A summary of the parameters associated with the economic costs of CALD suicide is provided in Table 2.

**Table 2.**
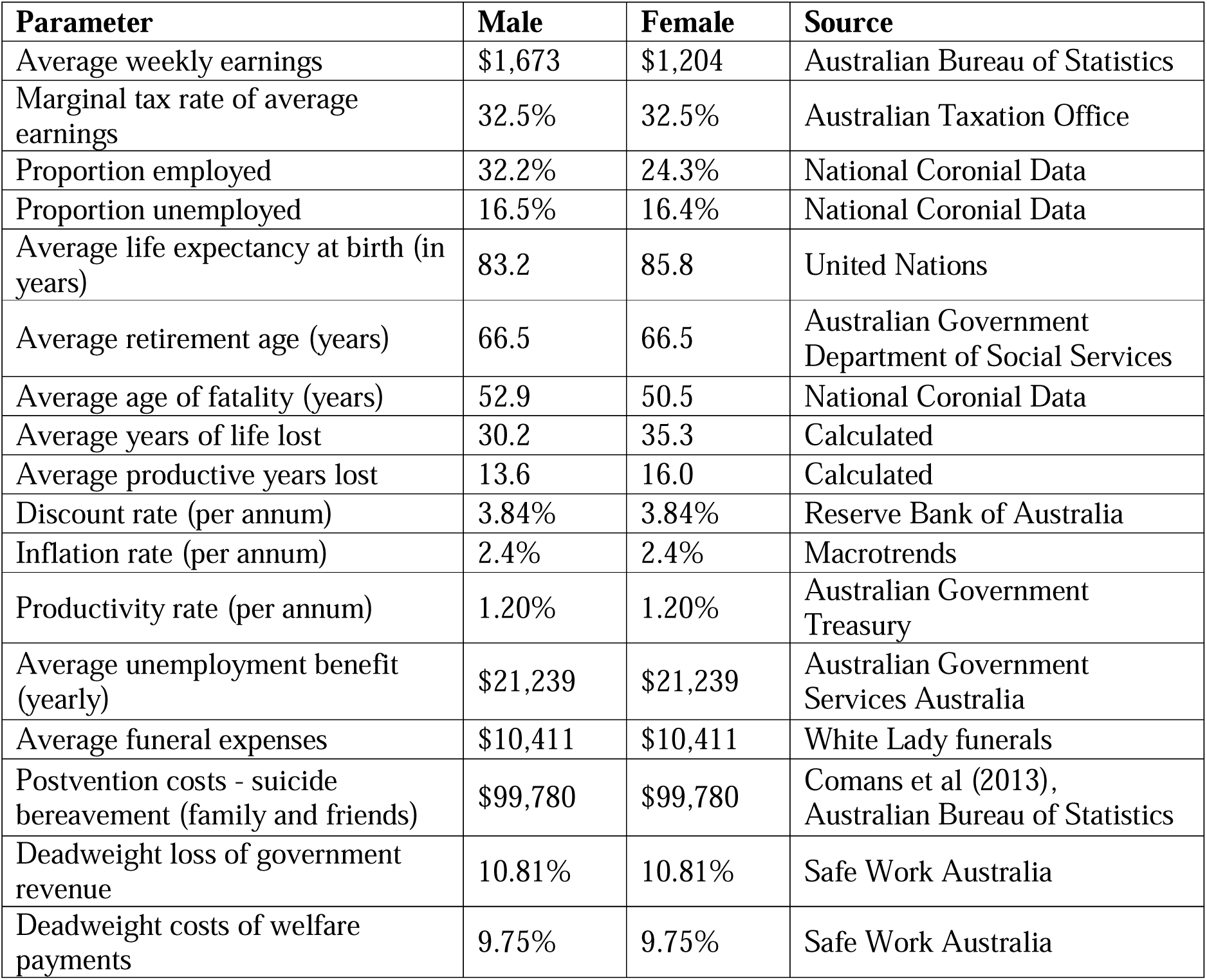
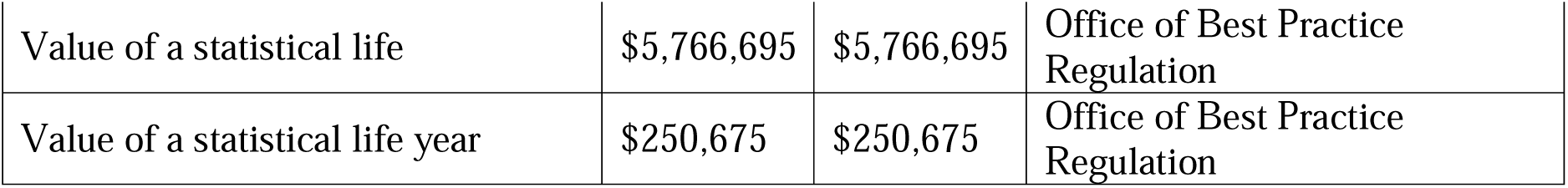
Summary of key cost parameters.

| Parameter | Male | Female | Source |
| --- | --- | --- | --- |
| Average weekly earnings | \$1,673 | \$1,204 | Australian Bureau of Statistics |
| Marginal tax rate of average earnings | 32.5% | 32.5% | Australian Taxation Office |
| Proportion employed | 32.2% | 24.3% | National Coronial Data |
| Proportion unemployed | 16.5% | 16.4% | National Coronial Data |
| Average life expectancy at birth (in years) | 83.2 | 85.8 | United Nations |
| Average retirement age (years) | 66.5 | 66.5 | Australian Government Department of Social Services |
| Average age of fatality (years) | 52.9 | 50.5 | National Coronial Data |
| Average years of life lost | 30.2 | 35.3 | Calculated |
| Average productive years lost | 13.6 | 16.0 | Calculated |
| Discount rate (per annum) | 3.84% | 3.84% | Reserve Bank of Australia |
| Inflation rate (per annum) | 2.4% | 2.4% | Macrotrends |
| Productivity rate (per annum) | 1.20% | 1.20% | Australian Government Treasury |
| Average unemployment benefit (yearly) | \$21,239 | \$21,239 | Australian Government Services Australia |
| Average funeral expenses | \$10,411 | \$10,411 | White Lady funerals |
| Postvention costs - suicide bereavement (family and friends) | \$99,780 | \$99,780 | Comans et al (2013), Australian Bureau of Statistics |
| Deadweight loss of government revenue | 10.81% | 10.81% | Safe Work Australia |
| Deadweight costs of welfare payments | 9.75% | 9.75% | Safe Work Australia |
| Value of a statistical life | \$5,766,695 | \$5,766,695 | Office of Best Practice Regulation |
| Value of a statistical life year | \$250,675 | \$250,675 | Office of Best Practice Regulation |

##### 3.1. Direct cost

Direct cost in this analysis considers funeral costs and postvention bereavement costs. Although there are many other potential direct costs, such as medical, first responder or coronial inquiry costs, evidence suggests that these costs account for a small proportion of total costs (Productivity Commission, 2020; Kinchin & Doran, 2017; Doran & Kinchin, 2020).

Funeral costs are estimated at $10,411(White Lady Funerals, 2023). It is acknowledged that funeral costs will vary by cultural or religious beliefs, so a conservative estimate is applied. Further, while funeral expenses may be associated with all deaths, fatality by suicide brings these costs forward. A fatality by suicide has a flow-on effect, with research suggesting that each fatality by suicide directly impacts between 6 and 135 people, including family members, work colleagues, friends, and first responders present at the time of death (Cerel *et al*., 2019; Comans, Visser & Scuffham, 2013). A conservative estimate is taken by assuming that the economic cost associated with suicide bereavement is estimated at $16,630 per person multiplied by six people bereaved (adjusted to reflect 2023 dollars) (Comans, Visser & Scuffham, 2013; Australian Bureau of Statistics, 2023).

##### 3.2. Indirect cost

Employment status by gender was obtained from NCIS. For those employed, lost economic productivity was calculated using the human capital method. This method considers the value of potential future earnings from the incident to the retirement age, assuming a discount profile and productivity loss. Average weekly earnings were estimated at $1,672.70 and $1,204.40 for males and females (Australian Bureau of Statistics, 2023), respectively. It is assumed that those who were unemployed received unemployment benefits, equivalent to an average allowance of $816.90 per fortnight (Services Australia, 2023). For those unemployed, the future income stream represents a saving to the Government. A long-term productivity factor of 1.2% per annum reflects growth in income/welfare (Treasury 2022). This figure is used in conjunction with a discount profile that includes a discount rate of 3.84% (average over the period 1990-2022) (Reserve Bank of Australia, 2023) adjusted for an inflation rate of 2.4% (average over the period 1990-2022) (Macrotrends, 2023) to determine the present value of future income streams. Also included in the indirect cost category are loss of government revenue through taxation and deadweight losses associated with the administration of taxation and welfare payments. Loss of government revenue reflects the tax losses due to foregone income and is valued using the marginal tax rate appropriate to the average weekly earnings (i.e., 32.5%) (Australian Taxation Office, 2023). Deadweight costs due to inefficiencies incurred through tax loss are estimated at 10.81% of the total net present value of loss of government revenue (i.e., taxation revenue). Deadweight costs due to inefficiencies incurred by social welfare payments are estimated at 9.75% of the total net present value of welfare payments (Safe Work Australia, 2015).

##### 3.3. Intangible cost

Intangible costs relate to the community value of a lost life cost that is estimated using a ‘willingness to pay’ approach based on the value of a statistical life. As noted in the Productivity Commission report (2020), the Bureau of Transport Infrastructure and Regional Economics (2011) uses this approach to calculate the costs associated with road fatalities. The value of a statistical life is an estimate of the financial value society places on reducing or avoiding the death of one person. By convention, it is assumed to be based on a healthy person living for another 40 years. It is known as a ‘statistical’ life because it is not the life of any particular person. An estimate of the value of life is, therefore, a tool for decision-making, not the value that is placed on any particular person. There are a variety of methods used to value a life, but the ‘willingness to pay’ method is viewed as the most appropriate technique (Office of Best Practice Regulation, 2019). Unlike other methods, such as the human capital model, which captures the discounted value of future earnings, the willingness to pay method quantifies non-market preferences and values, such as quality of life, health, and leisure (Australian Safety and Compensation Council. (2008). The Office of Best Practice Regulation (2019) has estimated the value of a statistical life year to be $250,675 million, adjusted to 2023 dollars.

## 3. Results

Table 3 provides an overview of CALD suicides in Australia in 2020. A total of 346 CALD Australians aged 15 and above died by suicide in 2020. Similar to the general population, male suicide was higher than females, with 70% of suicide deaths being of males. Women’s mean age of suicide was slightly lower than that of men. The age-standardised suicide rates of males were 11.6 per 100,000 and females were 5.1 per 100,000. Threat to breathing (caused by hanging) was the most common reason for suicide, both in males and females.

**Table 3.**
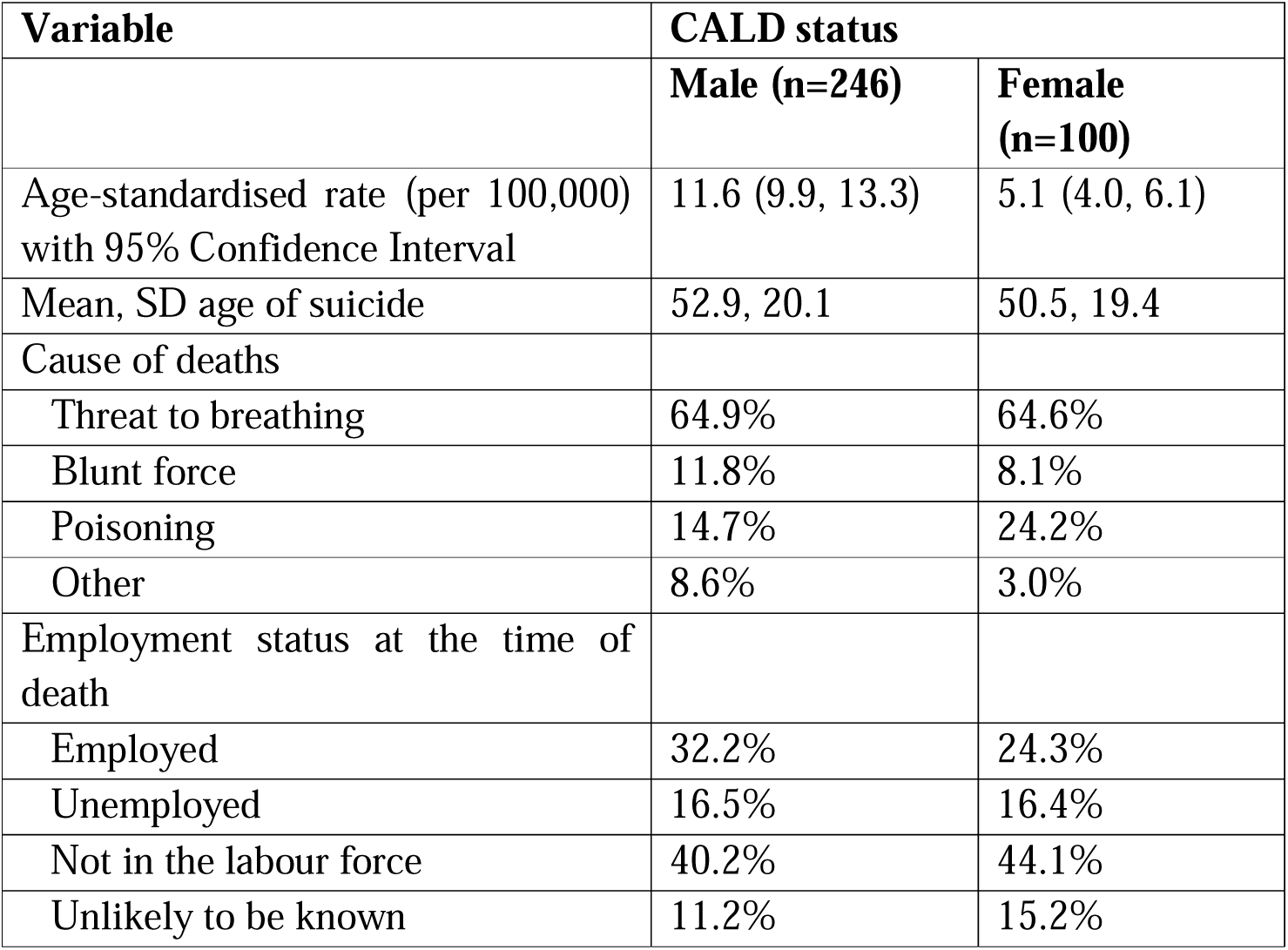
Profile of CALD suicide (15 and above) in Australia, 2020.

Table 4 provides an overview of the economic cost for the CALD population. Using data from the table, the average cost per fatality is $8.04 million and $9,23 million for males and females, respectively. Combining average cost with the number of fatalities results in a total economic cost associated with CALD suicide at $2,932 million (or $2.93 billion) each year. Applying the value of a statistical life year to the total number of potential years lost is the key cost driver in cost estimates, accounting for 94% of all costs. Although the average cost is higher in females than males, the total cost for males is estimated at $1,977 million compared with $923 million for females, driven by the larger number of suicides in males.

**Table 4:** Economic cost breakdown of CALD population in Australia.

|  | Male | Female | Total |
| --- | --- | --- | --- |
| CALD suicide deaths - 2020 | 246 | 100 | 346 |
| Potential years of life lost | 7,441 | 3,534 | 11,355 |
| Potential years of productive life lost | 3,340 | 1,600 | 5,116 |
| **Direct cost** |  |  |  |
| Total funeral expenses | \$2,561,212 | \$1,041,143 | \$3,602,355 |
| Total postvention costs | \$24,545,880 | \$9,978,000 | \$34,523,880 |
| **Sub total direct costs** | **\$27,107,092** | **\$11,019,143** | **\$38,126,235** |
| **Indirect costs** |  |  |  |
| Net present value of lost income for those employed (excluding taxation revenue lost) | \$64,274,815 | \$24,003,979 | \$119,225,928 |
| Net present value of taxation foregone | \$30,947,133 | \$7,801,293 | \$38,748,427 |
| Net present value of deadweight loss due to reduced government taxation | \$3,345,385 | \$843,320 | \$4,188,705 |
| Net present value of JobSeeker payments saved for those unemployed | -\$11,888,326 | -\$5,490,109 | -\$17,378,435 |
| Net present value of deadweight gain due to fewer JobSeeker payments | -\$1,159,112 | -\$535,286 | -\$1,694,397 |
| **Sub total indirect costs** | **\$85,519,895** | **\$26,623,198** | **\$143,090,227** |
| **Intangible costs** |  |  |  |
| Value of a statistical life year | \$250,675 | \$250,675 | \$250,675 |
| Total value of life lost | \$1,865,322,543 | \$885,769,399 | \$2,751,091,943 |
| ***Sub-total intangible costs*** | ***\$1,865,322,543*** | ***\$885,769,399*** | ***\$2,751,091,943*** |
| **TOTAL ECONOMIC COST** | **\$1,977,949,530** | **\$923,411,740** | **\$2,932,308,404** |

## Discussion

The estimated economic cost associated with suicide deaths among CALD individuals is $2.9 billion (in 2023 Australian dollars). This estimate is based on 346 deaths, representing approximately 10% of the total suicide deaths in Australia in 2020 (Australian Institute of Health and Welfare, 2023). These results are consistent with estimates generated by the Australian Government Productivity Commission (2020) at an average cost of $9.4 million (2018 AUD).

Our study is the first to demonstrate the economic impact of suicide among diverse populations not only in Australia but worldwide, where no such evidence is available for migrants or ethnic minorities. While we used economic tools and robust methods, we recognise the immeasurable loss of life and the emotional trauma experienced by the families and friends of those who died by suicide. Some migrants arrive in Australia with career aspirations, while others seek refuge from traumatic events, hoping for a better future. It is deeply concerning that some of those were unable to see a way forward and did not find the help they needed at the most distressful time of their lives.

Understanding the unique stressors faced by migrants with suicidal behaviours, coping mechanisms, and help-seeking behaviours in the event of distress is critically important for effective suicide prevention. Current evidence highlights some of the risk factors unique to the CALD population, including acculturation stress (Hovey, 2000), unemployment, and loneliness (Forte *et al*., 2018). Seeking help for suicidal thoughts or mental health conditions remains stigmatised for many migrants, and those who do seek help (Botchway-Commey et al., 2024; Wood & Newbold, 2012), do not always find it effective (Minhas, 2013; Dinham, 2020; Darmadi, 2022). Evidence indicates that suicide rates remain stagnant or increasing over time across different migrant groups in Australia (Maheen & King, 2023), with certain CALD groups exhibiting elevated suicide risk parameters (Maheen *et al*., 2024; Pham *et al*., 2023; Australian Institute of Health and Welfare, 2023).

In Australia, there have been some programs and projects on suicide prevention that target CALD populations. Notable programs include the translation of suicide prevention resources into multiple languages (Embrace Australia, 2020), translation and interpretation services at Primary Health Network’s commissioned mental health services, and programs for trauma victims from refugee backgrounds (Australian Centre for Health Services Innovation, 2022). There are also short-term projects, including gatekeeper training programs for faith leaders and other community members, training culturally diverse media outlets on responsible reporting of suicide incidents (Roses in Ocean, 2018), and training for Services Australia personnel (Queensland Transcultural Mental Health Centre, 2022). While these initiatives are significant, their focus is either limited to humanitarian entrants with trauma exposure or addressing language barriers, not the stressors that are associated with migrant suicide. Given the significant economic costs linked to CALD suicide, there is a strong rationale for developing a comprehensive national suicide prevention program. This program should address the multifaceted contributors to suicidal behaviours within these communities and be adaptable for implementation across diverse social, welfare, and health service settings.

### Limitations

The study utilised data from the NCIS, which reports only country of birth and usual country of residence to indicate migration status. This approach may underestimate individuals from CALD backgrounds who were born in Australia or in any English-speaking country and potentially include some Australians who were born overseas. The NCIS does not collect data on ethnicity or migration indicators, which could further underestimate the true representation of CALD populations. Despite these limitations, the country of birth remains a pivotal proxy indicator of migrant status widely employed in peer-reviewed literature and government reports. In Australia, given that previous migration waves have been linked with the countries of origin (migration from the UK, China, Europe, Asia, and the Middle East), the country of birth provides a useful migration context. The Australian Census and some population surveys collect a range of diversity indicators such as country of birth, parents’ country of birth, language spoken at home, language proficiency, years of migration, age at migration, or current migration status (Australian Bureau of Statistics, 2021; Watson & Wooden, 2021). We recommend the same of these be integrated into coroners’ reporting guidelines for better understanding the intersectionality of suicide within the CALD population, as well as measuring the social and economic impact of suicide among these groups.

It is important to note that 2020 was immediately after the onset of the COVID-19 pandemic. Fortunately, suicide rates in Australia remained unchanged during the pandemic, with slightly fewer suicide deaths reported compared to other years (Australian Institute of Health and Welfare, 2023); therefore, this should not affect the interpretation of our findings.

Authors of costing studies note that economic costing is not an exact science, and due to various limitations in data and methods, various assumptions are required (Kinchin & Doran, 2017; Doran & Kinchin, 2020; O’Dea *et al*., 2005, Kinchin & Doran, 2018; Doran *et al.,* 2016; ConNetica Conculting 2009). As noted by the Productivity Commission (2020), their estimates of the economic cost of suicidal behaviour are considered conservative. For example, their estimates exclude government expenditure directly on suicide prevention activities. The Australian Government spent almost $50 million on suicide prevention under its National Suicide Prevention Program in 2017. State and Territory Governments also fund their own suicide prevention activities designed to meet local needs. However, this expenditure is currently not being publicly reported or consistently consolidated.

It is also noteworthy that these estimates do not capture costs associated with suicidal ideation, thoughts or self-harm attempts. Recent data from the Australian National Study of Mental Health and Wellbeing suggest that for Australians aged 16-85 years, 16.7% had thoughts of suicide, 7.7% had made plans to suicide, and 4.8% had attempted suicide in their lifetime (Australian Bureau of Statistics, 2023). As noted by Kinchin *et al*. (2017), self-harming behaviour also results in a range of costs, including loss of potential years of life and productive years of life.

## Conclusion

Suicide and self-harm are significant global public health challenges with substantial economic implications. This study highlights the significant economic impacts of suicide among CALD populations and emphasises the need for a comprehensive national suicide prevention program tailored to diverse communities. Effective engagement with CALD communities is crucial for designing these initiatives, as it helps to understand how suicide manifests in different cultures, how help is sought, and which coping mechanisms are successful. Such engagement will enable the development of more efficient suicide prevention strategies, ultimately leading to a reduction in the economic costs associated with suicide and self-harm behaviours.

## Data Availability

All data produced in the present work are contained in the manuscript. Data used this study is not available on public request, and can only be obtained from the National Coroners Information System.

## Publication Ethics

The study was approved by the Justice Human Research Ethics Committee (reference, C.F./18/22468) and the Human Ethics Advisory Group (2022-23605-29220-3), School of Population and Global Health, University of Melbourne.

## Authorship

Humaira Maheen: Conceptualisation, original draft, writing, review and editing

Chris Doran: Data curation, writing, review and editing

All authors approved the final version of the article.

